# Normative Reference Values for the FACE-Q Skin Cancer Module: Patient Preoperative Scores and Comparison With Healthy Partners

**DOI:** 10.64898/2026.02.12.26345805

**Authors:** Maarten J. Ottenhof

## Abstract

**Background:** The FACE-Q Skin Cancer Module is a condition-specific patient-reported outcome measure for facial skin cancer. While its psychometric properties have been established, normative reference values that enable score interpretation in clinical practice and research are lacking.

**Objective:** To establish normative reference values for the FACE-Q Skin Cancer Module using preoperative patient data and to validate these values by comparison with a demographically matched cohort of healthy partners.

**Methods:** Two cohorts were analyzed: 287 patients with facial skin cancer (preoperative scores) and 82 healthy partners of skin cancer patients (same-age population without facial skin cancer). Both cohorts completed the Appearance (9 items) and Psychosocial Distress (8 items) scales. Patients additionally completed the Cancer Worry scale (10 items) and Sun Protection scale (5 items). Scores were transformed to a 0–100 scale. Normative values were expressed as percentiles overall and stratified by sex and age group. Group comparisons used independent t-tests, Mann–Whitney U tests, and Cohen’s d. Internal consistency was assessed with Cronbach’s alpha.

**Results:** Patient and partner cohorts were well matched for age (68.6±11.9 vs 68.4±13.0, p=0.902) and sex (46.7% vs 41.5% female, p=0.476). Surprisingly, preoperative facial appearance scores were virtually identical between patients and partners (55.6±14.0 vs 56.6±13.6, p=0.590, d=−0.08), as were psychosocial distress scores (14.3±12.0 vs 14.4±13.3, p=0.942, d=−0.01). This equivalence held across age groups. A significant sex interaction was identified: female patients scored lower on appearance than female partners (54.3 vs 59.9, p=0.048, d=−0.40), whereas no difference existed among males (56.9 vs 53.1, p=0.168). Internal consistency was excellent in both cohorts (Cronbach’s α 0.82–0.93). Patients reported marginally higher sun protection behaviors than partners (38.0 vs 33.6, p=0.050).

**Conclusions:** Preoperative FACE-Q Skin Cancer scores in patients are equivalent to those of demographically matched healthy individuals, confirming that these scores serve as valid normative references. The established percentile norms enable clinicians and researchers to interpret individual patient scores in context. The sex-specific difference in appearance scores warrants further investigation.

## Introduction

Patient-reported outcome measures (PROMs) are now standard tools for evaluating treatment quality and informing clinical decision-making. The FACE-Q Skin Cancer Module, developed using rigorous qualitative and psychometric methods, provides a condition-specific assessment of outcomes following facial skin cancer treatment. Its independently functioning scales address domains most important to patients: satisfaction with facial appearance, psychosocial distress, cancer-related worry, scar satisfaction, adverse effects of treatment, and sun protection behaviors.

While the development, validation, and cross-cultural adaptation of the FACE-Q Skin Cancer Module have been described, a fundamental tool for clinical application is still lacking: normative reference values. Without established norms, clinicians cannot determine whether an individual patient’s score is within the expected range, and researchers cannot contextualize group-level findings. Normative data are essential for identifying patients whose scores deviate meaningfully from the population average—for example, those with unusually low preoperative appearance satisfaction who may benefit from psychological support, or those with high cancer worry warranting targeted counseling.

Setting up norms for disease-specific questionnaires is tricky. Unlike generic health-related quality of life measures that can be normed on the general population, the FACE-Q Skin Cancer Module addresses constructs (e.g., cancer worry, scar satisfaction) that are inherently linked to the disease experience. We took two approaches: (1) using preoperative patient scores as condition-specific norms (reflecting the typical patient’s baseline), and (2) comparing with a healthy reference population to establish whether disease-related constructs are present even before treatment.

The present study aimed to establish normative reference values for the FACE-Q Skin Cancer Module through both approaches. We used preoperative data from 287 patients to generate percentile-based norms stratified by sex and age. Additionally, we compared these patient norms with scores from 82 healthy partners of skin cancer patients who completed the same instrument about their own faces, providing external validation of the normative framework.

## Methods

### Study Design and Population

This cross-sectional study analyzed data from two cohorts recruited at a tertiary academic medical center. The patient cohort comprised 287 consecutive patients with histologically confirmed facial skin cancer who completed the FACE-Q Skin Cancer Module preoperatively (before any surgical treatment). The partner cohort comprised 82 healthy partners (spouses or cohabiting partners) of skin cancer patients, who were invited to complete the same instrument about their own facial appearance and well-being. Partners were enrolled as a convenience sample; those with a personal history of facial skin cancer (n=4) or previous facial surgery (n=7) were not excluded but were flagged for sensitivity analysis. All participants provided informed consent. The study was approved by the institutional ethics committee.

### Outcome Measures

Both cohorts completed the Satisfaction with Facial Appearance scale (9 items, range 9–45) and Psychosocial Distress scale (8 items, range 8–40; higher scores indicate greater distress). Patients additionally completed the Cancer Worry scale (10 items, range 10–50; higher scores indicate more worry) and Sun Protection Behaviors scale (5 items, range 5–25). All raw sum scores were linearly transformed to a 0–100 scale for interpretability.

### Statistical Analysis

Demographic comparisons used independent t-tests for continuous variables and chi-squared tests for categorical variables. Score comparisons between patients and partners used independent t-tests, Mann–Whitney U tests, and Cohen’s d (pooled SD). Subgroup analyses stratified by sex and age group (<60, 60–70, >70 years) were performed. Internal consistency was evaluated using Cronbach’s alpha (≥0.70 considered acceptable, ≥0.80 good, ≥0.90 excellent). Normative reference values were expressed as mean±SD and as percentiles (5th, 10th, 25th, 50th, 75th, 90th, 95th) overall and stratified by sex and age. Analyses were performed in Python 3.11 using SciPy 1.11.

## Results

### Study Population

The patient cohort included 287 patients (mean age 68.6±11.9 years, range 18–93; 134 [46.7%] female). Fitzpatrick skin types were distributed as follows: type I, 48 (18.5%); type II, 102 (39.2%); type III, 106 (40.8%); types IV–V, 4 (1.5%). The partner cohort included 82 individuals (mean age 68.4±13.0 years, range 18–85; 34 [41.5%] female) with a similar skin type distribution (type I, 10 [16.1%]; type II, 19 [30.6%]; type III, 30 [48.4%]; type IV, 3 [4.8%]). There were no significant differences between patients and partners in age (p=0.902) or sex distribution (p=0.476).

### Patient–Partner Score Comparisons

Facial appearance satisfaction was virtually identical between patients and partners (55.6±14.0 vs 56.6±13.6, p=0.590, Cohen’s d=−0.08; Table 2). Similarly, psychosocial distress scores were indistinguishable (14.3±12.0 vs 14.4±13.3, p=0.942, d=−0.01). Both parametric and non-parametric tests confirmed the absence of significant differences (Mann–Whitney p=0.563 and p=0.904, respectively). This equivalence held across all three age groups (all p>0.05).

**Table 1.** Cohort Characteristics.

|  | Patients (n=287) | Partners (n=82) | p-value |
| --- | --- | --- | --- |
| Age, mean±SD | 68.6±11.9 | 68.4±13.0 | 0.902 |
| Female, n (%) | 134 (46.7%) | 34 (41.5%) | 0.476 |
| Skin type I, n (%) | 48 (18.5%) | 10 (16.1%) |  |
| Skin type II | 102 (39.2%) | 19 (30.6%) |  |
| Skin type III | 106 (40.8%) | 30 (48.4%) |  |

**Table 2.**
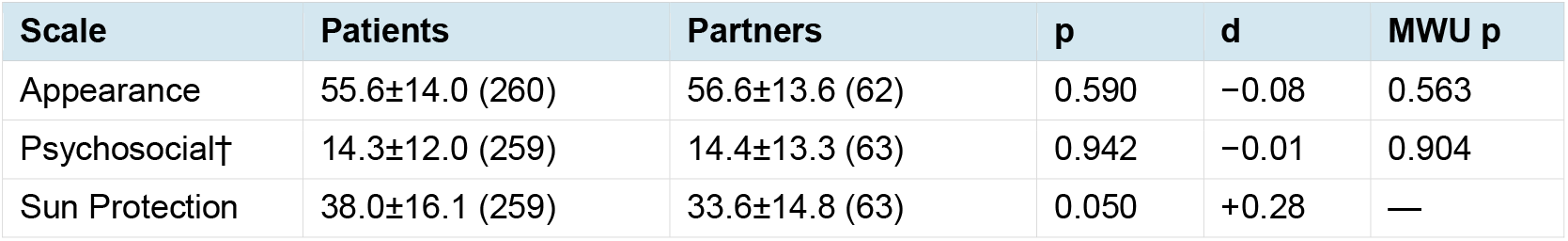
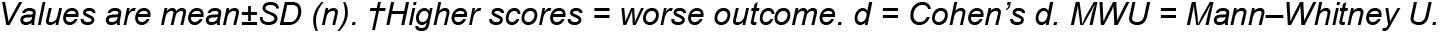
FACE-Q Score Comparisons (0–100 Scale)

| Scale | Patients | Partners | p | d | MWU p |
| --- | --- | --- | --- | --- | --- |
| Appearance | 55.6±14.0 (260) | 56.6±13.6 (62) | 0.590 | −0.08 | 0.563 |
| Psychosocial† | 14.3±12.0 (259) | 14.4±13.3 (63) | 0.942 | −0.01 | 0.904 |
| Sun Protection | 38.0±16.1 (259) | 33.6±14.8 (63) | 0.050 | +0.28 | — |

Values are mean ± SD (n). †Higher scores = worse outcome. d = Cohen's d. MWU = Mann–Whitney U.

**Table 3.** Normative Reference Values: Percentiles (0–100 Scale)

| Scale | n | Mean±SD | P5 | P10 | P25 | P50 | P75 | P90 | P95 |
| --- | --- | --- | --- | --- | --- | --- | --- | --- | --- |
| **Overall** |  |  |  |  |  |  |  |  |  |
| Appearance | 260 | 55.6±14.0 | 33.3 | 38.9 | 47.2 | 52.8 | 66.7 | 75.0 | 75.0 |
| Psychosocial† | 259 | 14.3±12.0 | 6.2 | 6.2 | 6.2 | 9.4 | 18.8 | 31.2 | 37.5 |
| Cancer Worry† | 259 | 25.2±14.7 | 0.0 | 7.0 | 15.0 | 25.0 | 32.5 | 45.5 | 52.5 |
| **Males** |  |  |  |  |  |  |  |  |  |
| Appearance | 126 | 56.9±13.5 | 33.3 | 38.9 | 47.9 | 52.8 | 68.8 | 75.0 | 77.8 |
| Psychosocial† | 125 | 13.2±11.4 | 6.2 | 6.2 | 6.2 | 9.4 | 15.6 | 31.2 | 37.5 |
| Cancer Worry† | 125 | 23.8±14.4 | 0.0 | 5.0 | 12.5 | 25.0 | 32.5 | 45.0 | 47.5 |
| **Females** |  |  |  |  |  |  |  |  |  |
| Appearance | 134 | 54.3±14.5 | 27.8 | 33.3 | 44.4 | 52.8 | 63.9 | 75.0 | 75.0 |
| Psychosocial† | 134 | 15.3±12.6 | 6.2 | 6.2 | 6.2 | 9.4 | 18.8 | 34.4 | 43.8 |
| Cancer Worry† | 134 | 26.5±15.0 | 0.0 | 7.5 | 15.0 | 25.0 | 34.4 | 50.0 | 55.0 |

†Higher scores = worse outcome. Values based on patient preoperative data.

However, a significant sex interaction emerged. Female patients scored significantly lower on facial appearance than female partners (54.3±14.5 vs 59.9±13.3, p=0.048, d=−0.40), suggesting that the anticipation of facial cancer surgery appears to hit harder on women’s appearance satisfaction. No such difference was observed among males (56.9±13.5 vs 53.1±13.2, p=0.168, d=0.28).

Patients reported marginally higher sun protection behaviors than partners (38.0±16.1 vs 33.6±14.8, p=0.050, d=0.28), which likely reflects increased awareness following skin cancer diagnosis.

### Internal Consistency

Internal consistency was excellent in both cohorts. For patients: Appearance α=0.928, Psychosocial α=0.820, Cancer Worry α=0.898. For partners: Appearance α=0.934, Psychosocial α=0.874. The comparable or higher reliability in the partner sample supports the validity of administering these scales to individuals without facial skin cancer.

## Discussion

This study establishes normative reference values for the FACE-Q Skin Cancer Module and validates them through comparison with a healthy partner cohort. The principal finding—that preoperative patient scores are virtually identical to those of age- and sex-matched healthy partners—has important implications for the interpretation and clinical application of this instrument.

The near-perfect equivalence between patients and partners on facial appearance satisfaction (d=−0.08) and psychosocial distress (d=−0.01) was unexpected and informative. We expected that patients diagnosed with facial skin cancer would report lower appearance satisfaction or greater distress preoperatively, reflecting anxiety about their diagnosis or the visible lesion itself. Surprisingly, a facial skin cancer lesion—typically a small basal or squamous cell carcinoma— does not substantially alter patients’ self-perceived facial appearance or psychological well-being compared to the general population. This may be because many facial skin cancers are relatively inconspicuous and grow slowly, giving patients time to adapt.

In practice, this means preoperative patient scores can serve as normative reference values: they represent the patient’s ‘healthy baseline’ and are equivalent to what would be obtained in a non-affected population. This makes interpreting scores much easier. A postoperative score below the 25th percentile of the normative distribution may signal a clinically relevant impairment; a score above the 75th percentile suggests an excellent outcome. The percentile tables provided in this study enable such comparisons for individual patients in routine clinical practice.

The sex-specific finding is striking. Female patients scored significantly lower on facial appearance than female partners (d=−0.40), whereas no difference existed among males. This small-to-medium effect size suggests that the prospect of facial cancer surgery appears to hit harder on women’s satisfaction with their appearance, consistent with broader literature on sex differences in body image and facial appearance concern. This finding points to the need for sex-stratified normative tables and may inform targeted preoperative counseling for female patients.

The marginally higher sun protection behaviors in patients compared to partners (p=0.050, d=0.28) is clinically intuitive. Patients with facial skin cancer have direct experience with the consequences of sun exposure and are likely to have received counseling about sun protection, which is reflected in their behavior. This difference, while modest, suggests that the FACE-Q Sun Protection scale is sensitive to clinically meaningful behavioral differences.

The excellent internal consistency observed in the partner cohort (Appearance α=0.93, Psychosocial α=0.87) validates administering FACE-Q Skin Cancer scales to individuals without the condition. The reliability was, if anything, slightly higher than in patients (Appearance α=0.93, Psychosocial α=0.82), suggesting that the items function at least as well in a normative population as in the target clinical population.

### Clinical Application

We propose the following framework for interpreting FACE-Q Skin Cancer scores using the normative data presented in this study. For facial appearance: scores below the 25th percentile (i.e., <47 on the 0–100 scale) indicate below-average satisfaction and may warrant clinical attention; scores above the 75th percentile (>67) indicate high satisfaction. For psychosocial distress: scores above the 75th percentile (>19) indicate above-average distress warranting further assessment; scores at or above the 90th percentile (>31) suggest clinically significant distress. For cancer worry: scores above the 75th percentile (>33) indicate substantial worry that may benefit from counseling; scores at the floor (0) indicate no worry, which was observed in 6.2% of patients preoperatively and may reflect either genuine absence of concern or response style effects.

### Limitations

Several limitations should be considered. First, the partner sample (n=82) is relatively small, particularly for stratified analyses. Second, partners were not randomly selected from the general population but recruited as convenience sample from the same clinical setting, which may introduce selection bias. Third, 4 partners (4.9%) had a personal history of skin cancer and 7 (8.5%) had undergone prior facial surgery; while flagged, excluding them would further reduce the sample size. Fourth, the partner sample only completed two of the five FACE-Q Skin Cancer scales (Appearance and Psychosocial), precluding normative comparison for Cancer Worry, Scars, and Adverse Effects. Fifth, the study was conducted at a single Dutch academic center; cross-cultural norms may differ. Finally, the normative tables are based on preoperative scores from a patient population (predominantly elderly, with facial skin cancer); they may not apply to populations with different demographic characteristics.

## Conclusions

This study provides the first normative reference values for the FACE-Q Skin Cancer Module. Preoperative patient scores are equivalent to those of healthy, demographically matched individuals, confirming their validity as normative benchmarks. The percentile tables enable clinicians and researchers to interpret individual scores in the context of the population distribution. Sex-stratified norms should be used when available, given the identified interaction between sex and patient/partner status for facial appearance satisfaction.

## Data Availability

The data underlying this study were collected at Clinique Rebelle, Amsterdam. Requests may be directed to the corresponding author. Clinic information: https://www.cliniquerebelle.com

